# Association of multidrug-resistant bacteria and clinical outcomes in patients with infected diabetic foot in a Peruvian Hospital: A retrospective cohort analysis

**DOI:** 10.1101/2024.02.12.24302725

**Authors:** Marlon Yovera-Aldana, Paola Sifuentes-Hermenegildo, Martha Sofia Cervera-Ocaña, Edward Mezones-Holguin

## Abstract

**Objective:** To evaluate the association of multidrug-resistant bacteria (MDRB) and adverse clinical outcomes in patients with infected diabetic foot (IDF) in a Peruvian hospital.

**Materials and Methods:** This retrospective cohort study evaluated patients treated in the Diabetic Foot Unit of a General Hospital in Lima, Peru. MDRB was based on resistance to more than two pharmacological groups across six clinically significant genera. The primary outcome was death due to complications of the IDF and/or major amputation. Other outcomes included minor amputation, hospitalization, and a hospital stay longer than 14 days. Relative risks were estimated using Poisson regression for all outcomes.

**Results:** The study included 192 IDF patients with a mean age of 59.9 years; 74% were males. A total of 80.8% exhibited MDRB. The primary outcome had an incidence rate of 23.2% and 5.4% in patients with and without MDRB, respectively (p = 0.01). After adjusting for sex, age, bone involvement, severe infection, ischemia, diabetes duration, and glycosylated hemoglobin, MDRB showed no association with the primary outcome (RR 3.29; 95% CI, 0.77 - 13.9), but did with hospitalization longer than 14 days (RR 1.43; 95% CI, 1.04 - 1.98).

**Conclusions:** Our study found no association between MDRB and increased mortality and/or major amputation but did find a correlation with prolonged hospitalization. The high proportion of MDRB could limit the demonstration of the relationship. It is urgent to apply continuous evaluation of bacterial resistance, implement a rational plan for antibiotic use, and maintain biosafety to confront this threat.

## Introduction

Infected Diabetic Foot (IDF) is one of the leading causes of hospitalization in diabetic foot units, with 15-20% of cases requiring major amputation[1,2]. In Peru, it accounts for 18.9% of diabetes-related hospital admissions, and 61% of these cases develop sepsis[3]. Treatment costs are five times higher for patients who develop this complication, which impacts the healthcare system and patients’ quality of life, particularly in developing countries[4]. Given the magnitude and significant impact of this complication, effective and personalized treatment is needed.

Antibiotic resistance poses a threat to the successful treatment of IDF[5]. The prevalence of Multidrug-Resistant Bacteria (MDRB) varies from 14% to 66%[6,7], depending on the country studied. MDRB is associated with protracted recovery, increased need for surgical procedures, prolonged hospital stays, and higher treatment costs[8]. Additionally, it results in increased protein-nutritional requirements, higher oxygen consumption, disturbed glycemic control, and reduced hemoglobin levels[9]. As these are polymicrobial infections, the use of broad-spectrum antibiotics is recommended for severe infections, subsequently deescalating according to the antibiogram[10]. For a proper IDF approach, it is mandatory to determine the bacteriological resistance profile at each healthcare facility and to devise a treatment algorithm based on those results[11].

The influence of MDRB on the clinical outcomes of IDF is not well understood[7], with scant research conducted in Latin America on the subject[3]. Factors such as inappropriate antibiotic selection due to the lack of an antibiogram, reduced antibiotic concentration in tissues caused by peripheral ischemia, and colonization by resistant strains due to frequent hospitalizations or prolonged treatments have been documented[12]. Furthermore, at the local level, only 30% of outpatient individuals have an HbA1c level below 7%, and a mere 10% have comprehensive metabolic control (involving lipids, weight, blood pressure, and blood glucose). Consequently, an adverse clinical outcome can potentially be expected when faced with an MDRB infection[13]. This study aims to assess the relationship between MDRB and its outcomes in IDF patients at a public hospital in Peru.

## Material and Methods

### Study Design and Setting

We conducted a retrospective cohort study on patients with DFI. We used secondary data from patients who attended the Diabetic Foot Unit (DFU) at the Maria Auxiliadora Hospital in Lima, Peru, between January 2017 and December 2019. This hospital is located in southern Lima and provides medical care to 2.5 million economically disadvantaged people. The DFU of the Endocrinology service has been the first line of treatment for IDF patients since 2015. Based on the severity, the DFU decides whether hospitalization or outpatient management is necessary.

### Patients

We included patients diagnosed with IDF as per the Infectious Diseases Society of America (IDSA) classification[10]. Additionally, those with a positive culture taken by the DFU staff within the first 48 hours of contact were included. Patients were excluded if they had extensive necrosis due to severe ischemia, no opportunity for culture, culture samples obtained via swabs or aspiration, follow-up of less than a month if not hospitalized, infected peripheral venous insufficiency, and pressure ulcers.

### Sample Size

After applying the inclusion and exclusion criteria, we obtained a sample size of 192 patients. The power of the sample was 88%, considering a proportion of 82% of the primary outcome in exposed subjects, 78% in non-exposed subjects, a confidence level of 95%, and a 4:1 ratio between exposed and non-exposed subjects.

### Variables

#### Multidrug-Resistant Bacteria (MDRB)

MDRB was defined as a lack of susceptibility to at least one agent in three or more classes of antimicrobials for each bacterial genus: *Staphylococcus sp*., *Enterococcus sp*., *Enterobacteriaceae*, *Pseudomonas sp*., and *Acinetobacter sp*. Innate resistances to some drugs were not considered for this definition. **S1-S5 Tables**. show the criteria to consider resistance for each bacterial genus[14].

#### Adverse Clinical Outcomes

The primary composite outcome consisted of death due to IDF complications and/or major amputation (above the ankle). Secondary outcomes were death related to the diabetic foot, major amputation, minor amputation (below the ankle), hospitalization solely for medical treatment, and prolonged stay (more than 14 days).

#### Diabetic Foot Characteristics

We described the characteristics of the lesion and the patient upon admission to the DFU using the San Elian scale. This tool includes 10 variables: description of depth, infection, ischemia, neuropathy, location, topography, number of zones, edema, and degree of inflammation[15]. The largest and smallest diameters of the ulcer were recorded, and the area was calculated using the ellipse formula. The degree of infection was based on the IDSA consensus[10]. Peripheral arterial disease classification was obtained from arterial plethysmography reports, with positive results if biphasic, monophasic, or absence of wave patterns were present. Peripheral neuropathy was diagnosed if there was a loss of protective sensation evaluated with a monofilament. The glomerular filtration was calculated using the CKD-EPI formula[16]. Previous exposure to antibiotics in the last year, as well as previous amputation or hospitalization for diabetic foot, was documented. Furthermore, we recorded the levels of hemoglobin, albumin, leukocytes, and creatinine at the time of the acute episode. We accepted HbA1c values up to one month old (**S6 Table**).

### Procedures

Authorization was obtained from the Endocrinology department head to access DFU data. The endocrinology service continuously records the outcomes of its patients for administrative purposes. This spreadsheet was provided without personally identifiable information. A first request was made in February 2019 and then in December 2019. Two authors completed other data of interest by reviewing the medical records from February 2019 to January 2020 if necessary. **(S7 Table)**.

#### Management Guide

The endocrinology service staff applied diagnostic and therapeutic procedures following the International Working Group of Diabetic Foot Guide for the management of infection, peripheral arterial disease, and ulcers[17]. The team consisted of four doctors and two nurses trained in comprehensive management.

#### Microbiological Analysis

The DFU staff obtained culture samples via tissue biopsy following the wound debridement using the standard procedure[18]. The hospital laboratory staff conducted the bacterial identification analysis and antibiotic susceptibility tests using an automated system, VITEK® 2 (BioMérieux Laboratory, Argentina)[19].

### Statistical Analysis

Clinical and laboratory characteristics were described quantitatively, using either mean with standard deviation or median with interquartile range, depending on the normality assessed by the Shapiro-Wilk test. Categorical variables were presented with absolute and relative frequencies. Moreover, the isolated bacteria were characterized by percentages, according to the Gram stain type and genus. To determine the differences between the groups with and without MDRB, we used the Student’s T-test or U-Mann Whitney test for numerical variables based on distribution. For categorical variables, we employed the Pearson’s Chi-square test or Fisher’s exact test. We also assessed the incidence of the primary composite outcome based on demographic and clinical characteristics.

A multivariate analysis was performed using a generalized linear model with a Poisson function, logarithmic link, and robust variance to determine the relative risk (RR) for the primary and secondary outcomes. An unadjusted model and three adjusted models were developed based on epidemiological variables. Model 1 was adjusted for age and sex, Model 2 for bone involvement, severe infection, and peripheral arterial disease, and Model 3 for diabetes duration and HbA1C levels. The analysis was conducted using the STATA® software (version 15.1, Texas, USA), and a significance level of 5% was applied for all hypothesis tests.

#### Ethics

The Institutional Ethics Committee for Research at María Auxiliadora Hospital approved the development of this research project, under the code HA/CIEI/019/19. The data provided by the Endocrinology Service did not contain any personally identifiable information. If it was necessary to review medical records for missing data, the collection forms did not contain identifying data. Only the lead author had access to the data.

## RESULTS

The initial database comprised 582 subjects evaluated by the UPD between 2017 and 2019. Based on eligibility criteria, we excluded 390 patients, with the primary reason for exclusion being the absence of a bacterial culture. Ultimately, 192 patients were included in the study (**Fig 1**).

**Figure 1.**
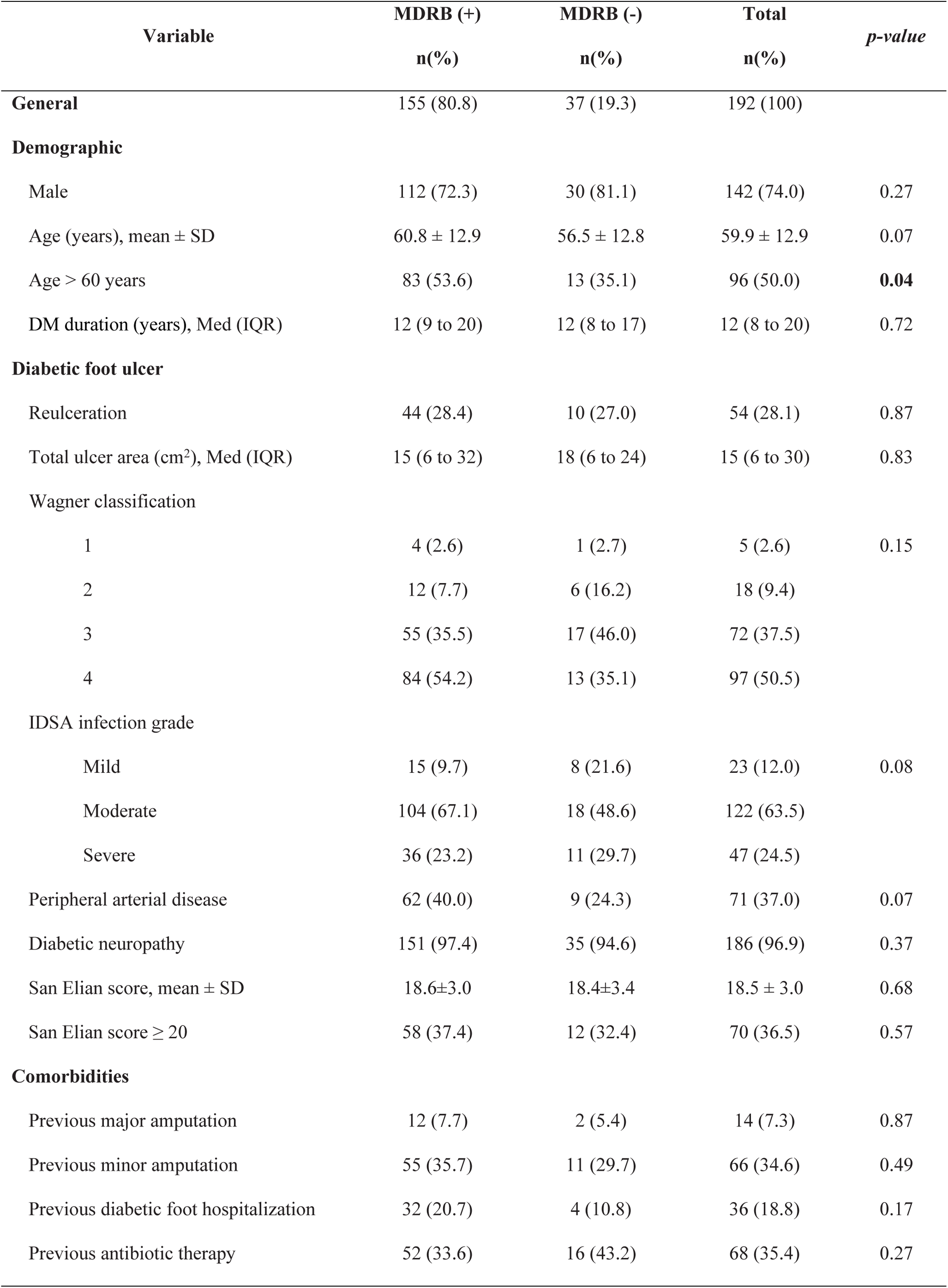
Flowchart of patients included in the study.

Out of the total, 155 patients (80.8%) had a MDRB infection, 74% were males, with an average age of 59.9 ± 12.9 years. The median duration of diabetes mellitus was 12 years, the average HbA1c was 10%, and 81% of the patients had an HbA1c level of ≥7. Notably, 28.1% of the patients developed a recurrent ulcer. As per the Wagner classification of DFI, 50.5% were classified as grade IV, and 88% had a moderate to severe infection according to the IDSA guidelines. The remaining demographic and clinical characteristics of the patients are shown in **Table 1**.

**Table 1.**
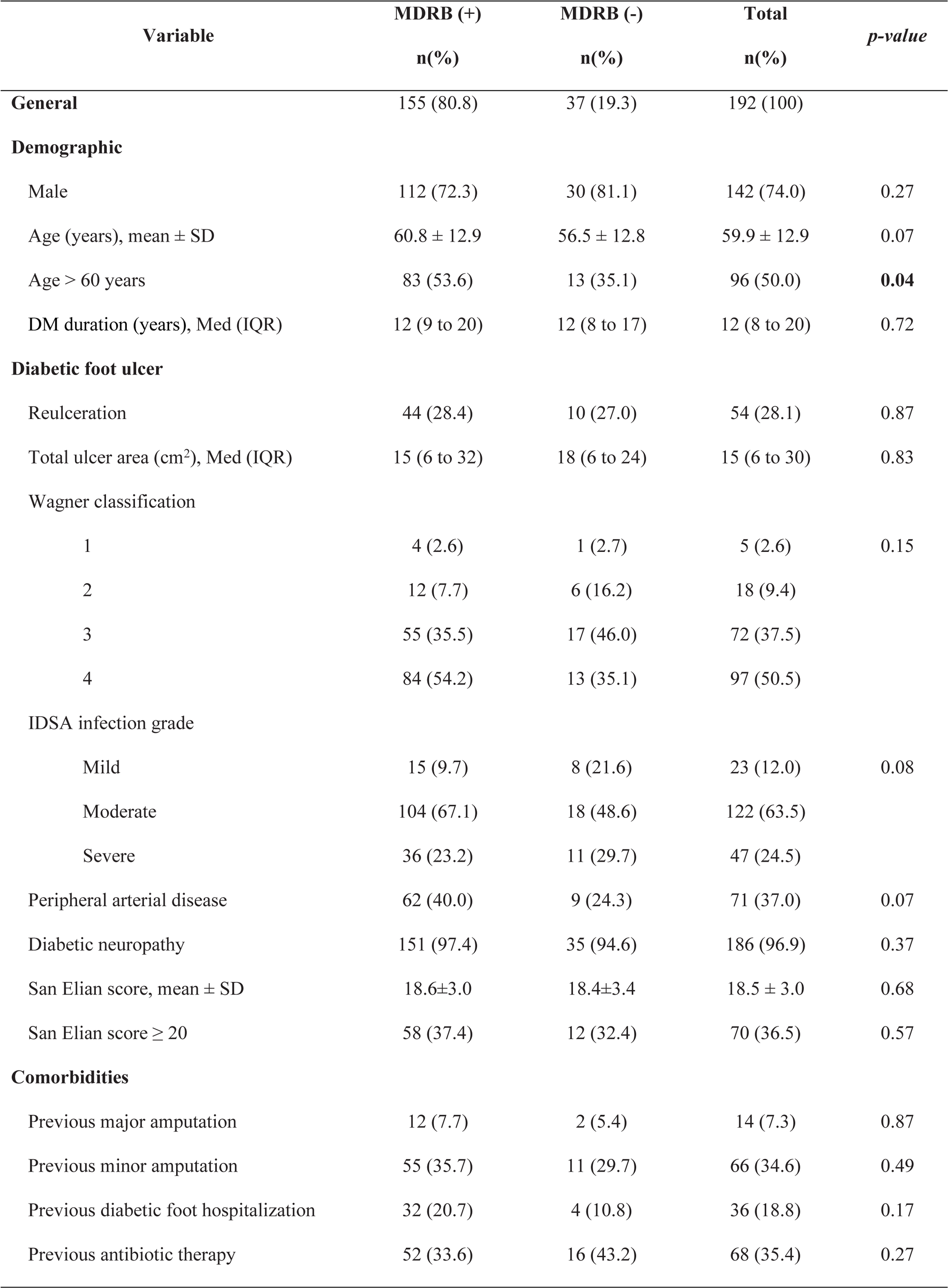
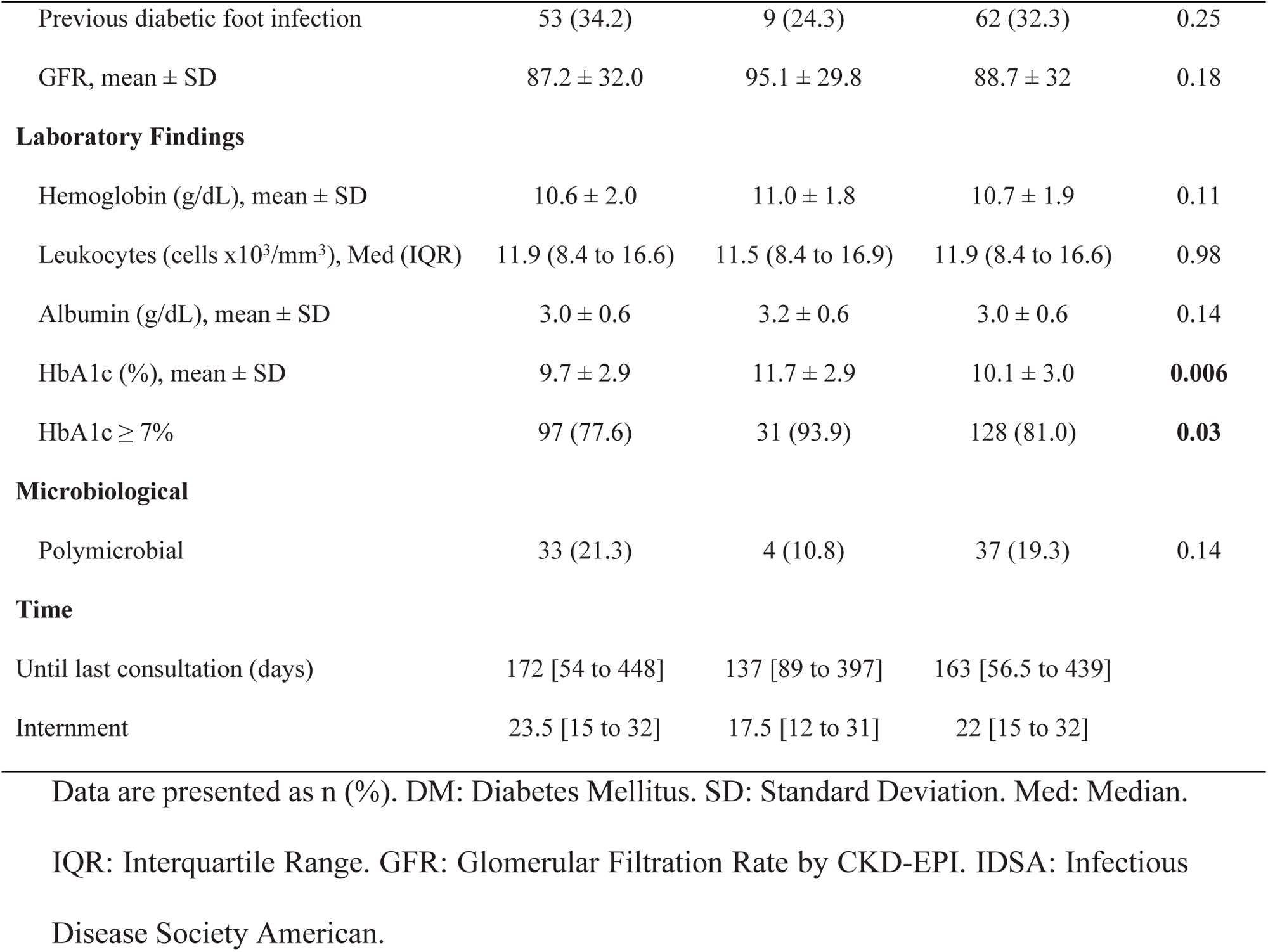
Clinical and demographic characteristics according to the MDR organism.

Of a total of 236 isolated bacteria, 82% were MDR. The most common MDR bacteria were *Enterobacteriaceae* (51.3%), *Staphylococcus aureus* (28.4%), and *Enterococcus sp.* (8.9%). Gram-negative bacteria represented 61% of the cases, and only 37 (19.3%) had a polymicrobial culture. A detailed description of the isolated MDR organisms is presented in **Table 2**.

**Table 2.**
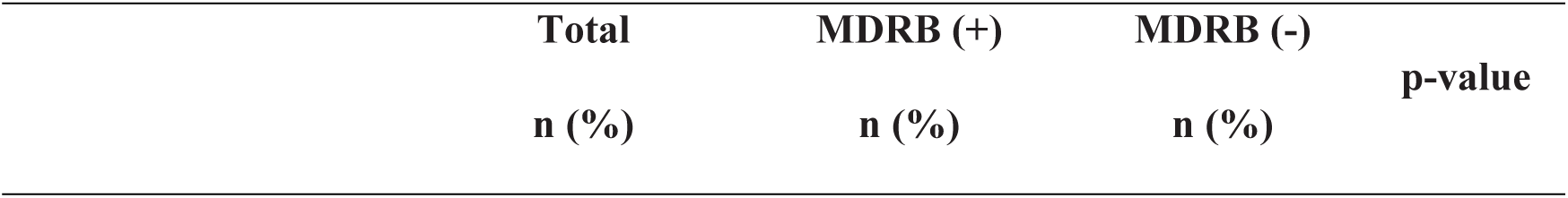
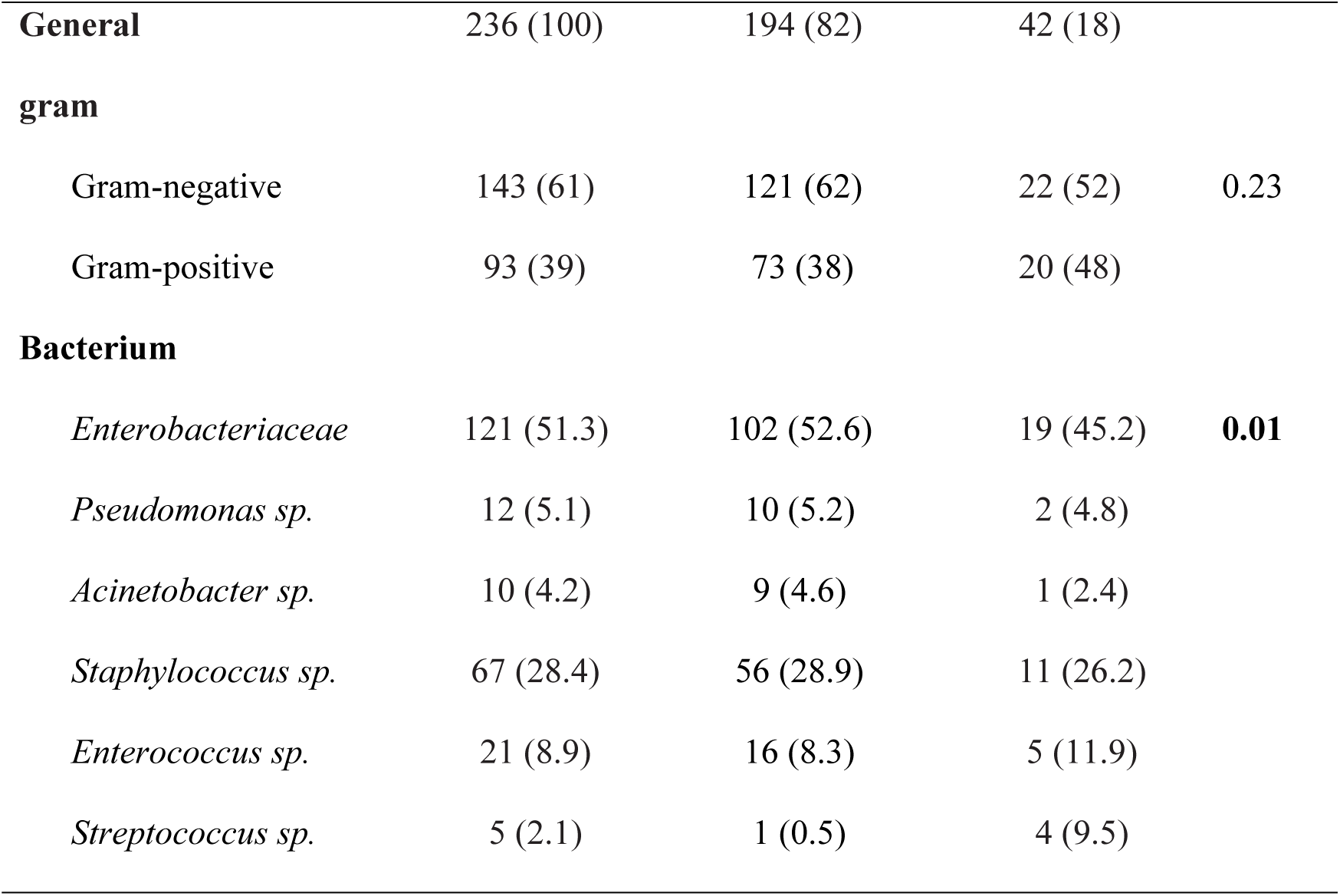
Microbiological profile of the isolated bacterium according to MDRB.

The primary outcome (death and/or major amputation) occurred in 18.9% of all patients. The mortality rate was 2.1%, and major amputation was observed in 17.7%. Regarding secondary outcomes, 67% of the general population required hospitalization. Of those hospitalized, 80% had a stay longer than 14 days (**Table 3 and S1 Fig)**.

**Table 3.** Clinical outcomes according to the MDRB pre-secretion.

|  | **Total** | **MDRB (+)** | **MDRB (-)** | **p-value** |
| --- | --- | --- | --- | --- |
|  |  | **n (%)** | **n (%)** |  |
| **Composite primary outcome** | 38 /192 (18.9) | 36/155 (23.2) | 2/37 (5.4) | **0.01** |
| **(n=192)** |  |  |  |  |
| **Secondary outcomes** |  |  |  |  |
| Mortality (n=192) | 4/192 (2.1) | 3/155 (1.9) | 1/37 (2.7) | 0.77 |
| Major amputation (n=188) | 34/188 (17.7) | 33/152 (21.7) | 1/36 (2.8) | **0.008** |
| Minor amputation<sup>b</sup> (n=154) | 82/154 (53.2) | 65/119 (54.6) | 17/35 (48.6) | 0.53 |
| Hospitalization<sup>b</sup> (n=154) | 90/154 (67.0) | 66/119 (55.5) | 24/35 (68.6) | 0.17 |
| Long stay<sup>c</sup> (n=90) | 66/90 (80.0) | 53/68 (80.3) | 13/24 (54.2) | **0.013** |

<sup>b</sup>Does not include fatalities or major amputations. <sup>c</sup>Only in hospitalized.

The demographic characteristics associated with the primary composite outcome were age over 60 years, duration of diabetes over 10 years, ulcer area larger than 10 cm^2^, depth of the lesion according to Wagner classification[20], infection grade according to IDSA[10], presence of peripheral arterial disease, San Elian score ≥ 20[15], hemoglobin <10 g/dl, albumin <3.5 g/dl, HbA1c ≥7%, and polymicrobial culture (**Table 4**).

**Table 4.**
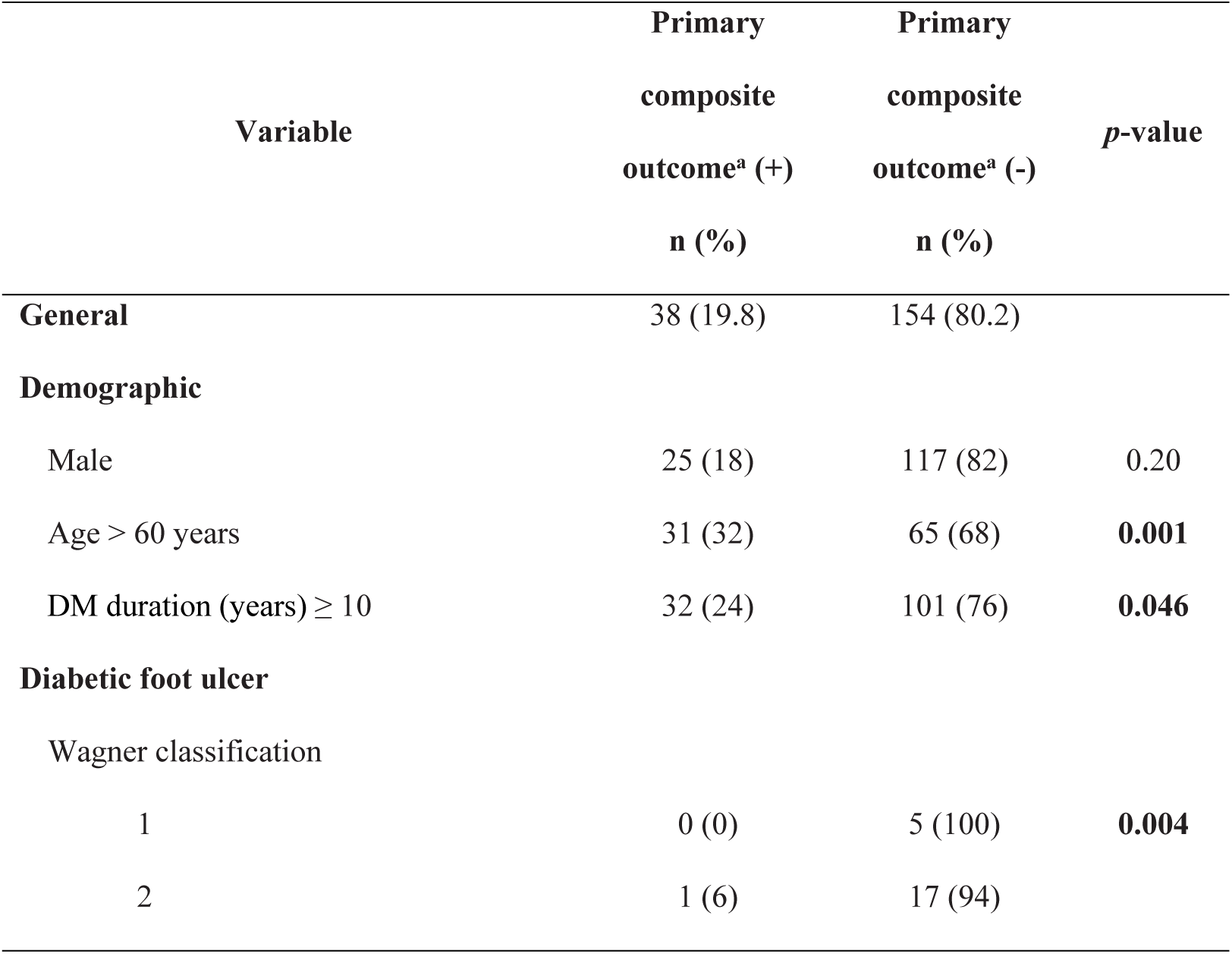
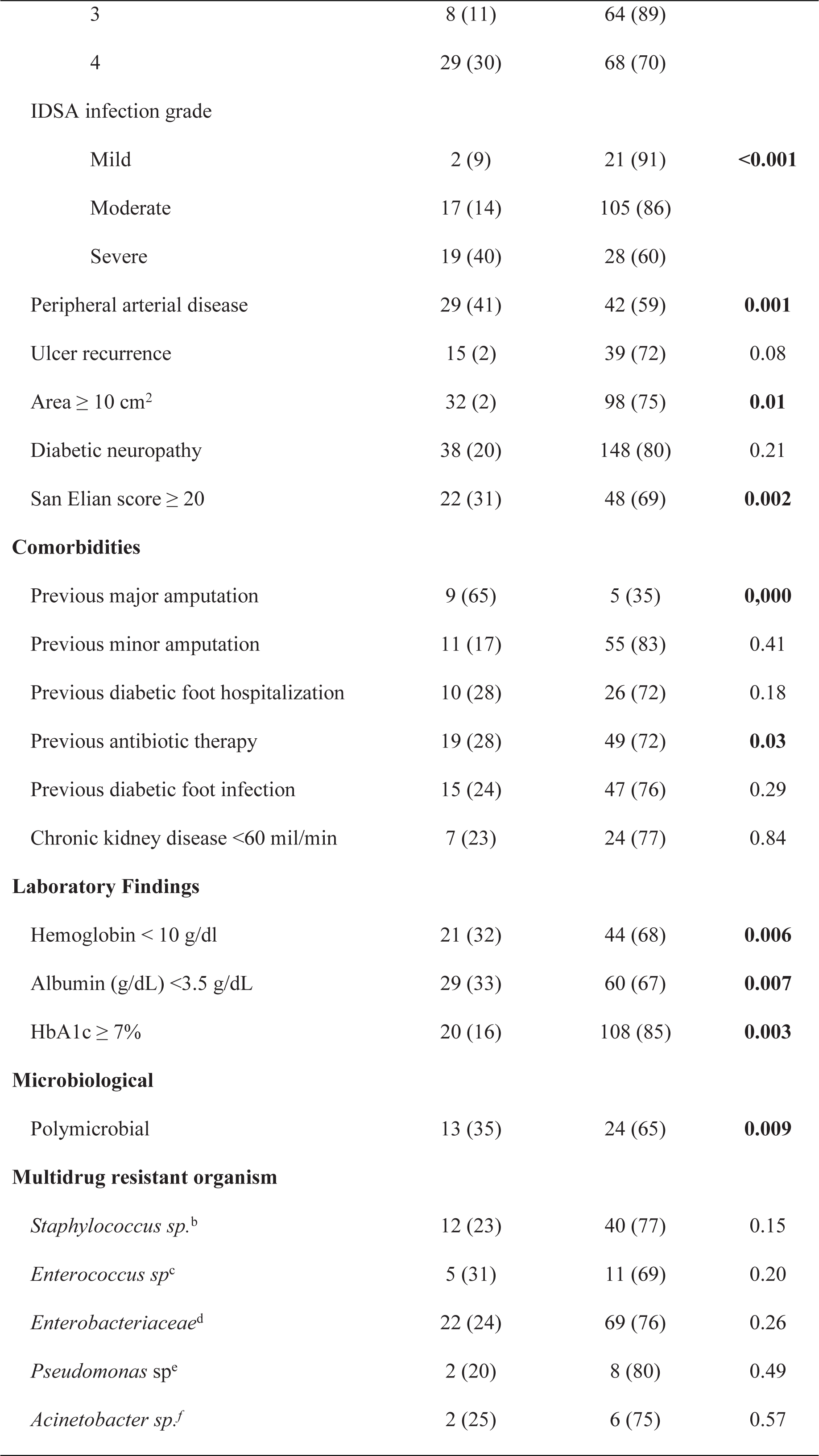
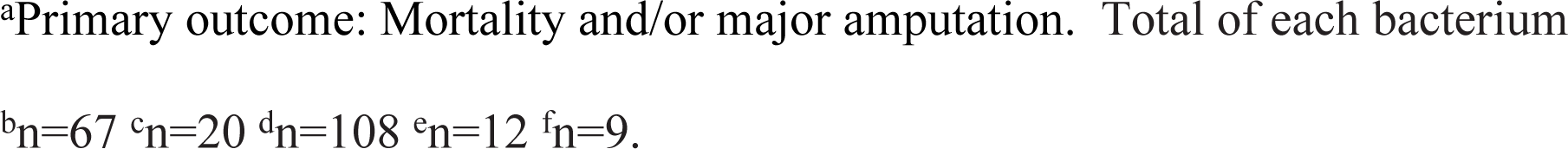
Association between primary composite outcome and population characteristics.

| Variable | Primary<br>composite<br>outcome<sup>a</sup> (+)<br>n (%) | Primary<br>composite<br>outcome<sup>a</sup> (-)<br>n (%) | *p*-value |
| --- | --- | --- | --- |
| **General** | 38 (19.8) | 154 (80.2) |  |
| **Demographic** |  |  |  |
| Male | 25 (18) | 117 (82) | 0.20 |
| Age > 60 years | 31 (32) | 65 (68) | **0.001** |
| DM duration (years) $\geq 10$ | 32 (24) | 101 (76) | **0.046** |
| **Diabetic foot ulcer** |  |  |  |
| Wagner classification |  |  |  |
| 1 | 0 (0) | 5 (100) | **0.004** |
| 2 | 1 (6) | 17 (94) |  |
| 3 | 8 (11) | 64 (89) |  |
| 4 | 29 (30) | 68 (70) |  |
| IDSA infection grade |  |  |  |
| Mild | 2 (9) | 21 (91) | **<0.001** |
| Moderate | 17 (14) | 105 (86) |  |
| Severe | 19 (40) | 28 (60) |  |
| Peripheral arterial disease | 29 (41) | 42 (59) | **0.001** |
| Ulcer recurrence | 15 (2) | 39 (72) | 0.08 |
| Area $\geq 10$ cm<sup>2</sup> | 32 (2) | 98 (75) | **0.01** |
| Diabetic neuropathy | 38 (20) | 148 (80) | 0.21 |
| San Elian score $\geq 20$ | 22 (31) | 48 (69) | **0.002** |
| **Comorbidities** |  |  |  |
| Previous major amputation | 9 (65) | 5 (35) | **0.000** |
| Previous minor amputation | 11 (17) | 55 (83) | 0.41 |
| Previous diabetic foot hospitalization | 10 (28) | 26 (72) | 0.18 |
| Previous antibiotic therapy | 19 (28) | 49 (72) | **0.03** |
| Previous diabetic foot infection | 15 (24) | 47 (76) | 0.29 |
| Chronic kidney disease <60 ml/min | 7 (23) | 24 (77) | 0.84 |
| **Laboratory Findings** |  |  |  |
| Hemoglobin < 10 g/dl | 21 (32) | 44 (68) | **0.006** |
| Albumin (g/dL) <3.5 g/dL | 29 (33) | 60 (67) | **0.007** |
| HbA1c $\geq 7\%$ | 20 (16) | 108 (85) | **0.003** |
| **Microbiological** |  |  |  |
| Polymicrobial | 13 (35) | 24 (65) | **0.009** |
| **Multidrug resistant organism** |  |  |  |
| *Staphylococcus* sp.<sup>b</sup> | 12 (23) | 40 (77) | 0.15 |
| *Enterococcus* sp.<sup>c</sup> | 5 (31) | 11 (69) | 0.20 |
| *Enterobacteriaceae*<sup>d</sup> | 22 (24) | 69 (76) | 0.26 |
| *Pseudomonas* sp.<sup>e</sup> | 2 (20) | 8 (80) | 0.49 |
| *Acinetobacter* sp.<sup>f</sup> | 2 (25) | 6 (75) | 0.57 |

<sup>a</sup>Primary outcome: Mortality and/or major amputation. Total of each bacterium
<sup>b</sup><sub>n=67</sub> <sup>c</sup><sub>n=20</sub> <sup>d</sup><sub>n=108</sub> <sup>e</sup><sub>n=12</sub> <sup>f</sup><sub>n=9</sub>.

In the crude model, for patients with MDRB, the incidence of the primary composite clinical outcome increased by 3.3 times. However, when adjusted for epidemiological variables, no association was evidenced in any of the three proposed models. Among the secondary outcomes, only a hospital stay longer than 14 days was associated with MDRB when adjusted in any of the three proposed models. The outcomes and their adjusted RR can be observed in **Table 5**.

**Table 5.** Association between MDRB and adverse clinical events: crude and adjusted models.

|  | Model | Crude | *p*-<br>value | Model 1* | Adjusted | *p*-<br>value | Model<br>2** | Adjusted | *p*-value | Model<br>3*** | Adjusted | *p*-<br>value |
| --- | --- | --- | --- | --- | --- | --- | --- | --- | --- | --- | --- | --- |
|  | RR | 95% CI |  | RR | 95% CI |  | RR | 95% CI |  | RR | 95% CI |  |
| **Composite primary outcome<sup>a</sup>** |  |  |  |  |  |  |  |  |  |  |  |  |
| Not MDRB | 1.00 |  |  | 1.00 |  |  | 1.00 |  |  | 1.00 |  |  |
| MDRB | 4.29 | 1.08 – 17.1 | 0.039 | 3.40 | 0.86 – 13.4 | 0.081 | 3.28 | 0.99 – 10.8 | 0.051 | 3.29 | 0.77 – 13.9 | 0.105 |
| **Secondary outcomes** |  |  |  |  |  |  |  |  |  |  |  |  |
| **Mortality** |  |  |  |  |  |  |  |  |  |  |  |  |
| Not MDRB | 1.00 |  |  | 1.00 |  |  |  |  |  |  |  |  |
| MDRB | 0.71 | 0.08-6.72 | 0.770 | 0.27 | 0.04-1.96 | 0.194 | 0.53 | 0.05 – 5.91 | 0.611 | 0.27 | 0.05 – 1.66 | 0.158 |
| **Major amputation** |  |  |  |  |  |  |  |  |  |  |  |  |
| Not MDRB | 1.00 |  |  | 1.00 |  |  | 1.00 |  |  | 1.00 |  |  |
| MDRB | 7.81 | 1.1 – 55.5 | 0.040 | 6.45 | 0.91 – 45.4 | 0.061 | 5.41 | 0.88 – 33.2 | 0.068 | 6.55 | 0.89 – 47.9 | 0.064 |
| **Minor amputation<sup>b</sup>** |  |  |  |  |  |  |  |  |  |  |  |  |
| Not MDRB | 1.00 |  |  | 1.00 |  |  | 1.00 |  |  | 1.00 |  |  |
| MDRB | 1.12 | 0.76-1.64 | 0.544 | 1.15 | 0.78-1.69 | 0.474 | 0.98 | 0.68-1.40 | 0.908 | 1.20 | 0.81 – 1.77 | 0.350 |
| **Hospitalization<sup>b</sup>** |  |  |  |  |  |  |  |  |  |  |  |  |
| Not MDRB | 1.00 |  |  | 1.00 |  |  | 1.00 |  |  | 1.00 |  |  |
| MDRB | 0.81 | 0.61-1.07 | 0.133 | 0.84 | 0.64-1.11 | 0.222 | 0.77 | 0.59 – 1.01 | 0.059 | 0.81 | 0.59 – 1.10 | 0.182 |
| **Prolonged stay<sup>c</sup>** |  |  |  |  |  |  |  |  |  |  |  |  |
| Not MDRB | 1.00 |  |  | 1.00 |  |  | 1.00 |  |  | 1.00 |  |  |
| MDRB | 1.49 | 1.06 – 2.10 | 0.020 | 1.45 | 1.05 – 2.02 | 0.026 | 1.46 | 1.06 – 2.02 | 0.021 | 1.51 | 1.06 – 2.14 | 0.021 |

<sup>a</sup>Primary composite outcome: Mortality and/or major amputation. <sup>b</sup>Does not include deaths or major amputation ( $n=154$ ). <sup>c</sup>Only in hospitalized $n=90$ . RR: Relative risk. CI: confidence interval. \*Model 1: Adjusted for age and gender. \*\*Model 2: Adjusted for bone compromise, severe infection, and peripheral arterial disease. \*\*\* Model 3: Adjusted for diabetes duration and HbA1C.

## DISCUSSION

### Main Results

In this study, we found no association between MDRB and the primary outcome of death related to diabetic foot and/or major amputation, but we did find an association with a hospital stay longer than 14 days.

### Comparison with other studies

Different studies have shown controversial results regarding the influence of MDRB on the clinical course of DFI. Gupta et al.[7] and Richard et al.[21] concluded that MDRB is not associated with the clinical outcome of patients with DFI. In these studies, the lack of association was attributed to early aggressive treatment, antibiotic therapy adjusted to microbiological findings, and the small number of patients with MDRB included. In our study, after multivariate analysis, no association was defined between MDRB and a higher risk of major amputation. However, Matta et al.’s systematic review did find a higher prevalence of amputation in DFI with MDRB. However, these studies only performed univariate analysis and considered that the higher risk of amputation in their population is more attributed to the metabolic status or immunosuppression of the patients[22].

In our study, we observed an MDRB prevalence of 82% [9], a figure that is significantly higher compared to other countries such as France, India, China, and Turkey, where it fluctuated between 13.8 to 72.5%[22]. One notable outlier was Ethiopia, where a staggering 93% prevalence was reported[23]. This high rate of MDRB in our research might be attributable to several factors. We did not exclude patients who had received prior antibiotic treatment, a practice known to foster the emergence of resistant bacterial strains[21]. In addition, 20% of our patients had a history of hospitalization due to DFI, potentially predisposing them to selective colonization by resistant pathogens[24]. Furthermore, it’s essential to bear in mind that the definition of MDRB can vary across different studies, leading to discrepancies in reported prevalence rates based on the specific criteria employed.

The scientific literature suggests a difference in the prevalence of gram-positive or negative germs depending on the economic level. It is described that the prevalence of gram-negative bacteria is higher in developing countries, as in the studies by Gadepalli et al.[1] and Datta et al.[25]. Similarly, in our study, gram-negative bacteria were the most frequent, finding an 85% bone involvement and an 80% moderate-severe infection, which is related to infection by gram-negative or mixed bacteria. Also, the *Enterobacteriaceae* family presented the highest proportion of MDRB (52.6%), followed by the *Staphylococcus sp.* genus (28.9%), unlike the study by McDonald et al. [26], where *Staphylococcus sp.* was the most common MDR bacteria. The similarities of our results with the studies from India mentioned above[1,25], could be related to the similar degree of hygiene, health education, footwear use, and geographical area related to warm climates. It could also be related to the low budget that the health system invests to create programs that make the management and treatment of patients with DFI more efficient.

Prolonged hospitalization was the only adverse clinical event associated with MDRB, a finding similar to previous studies[8,24], which could suggest nosocomial acquisition through cross-transmission via caregivers’ hands[27,28]. The search for hospital care in these patients, as well as the extended stay, may be influenced by factors such as the complexity of the procedures, the longer duration of ulcers requiring prolonged antibiotic treatment, timely follow-up, and slow recovery of patients with MDRB[8]. Furthermore, it is worth highlighting that 50.5% of patients had a Wagner classification of 4 and 88% had a moderate-severe infection according to IDSA guideline, so it was expected that a longer hospitalization would be due to the severity of their clinical condition[29].

Other factors that modify the relationship between the presence of MDRB and adverse outcomes are polymicrobial culture, vascular insufficiency, anemia, and hyperglycemia. Polymicrobial culture has been associated with the severity of DFI. The symbiosis between two bacteria can potentiate the damage when they are individually present[30]. It is also suggested that vascular insufficiency leads to tissue hypoxia and also causes a decrease in antibiotic concentrations at the site[31]. Similarly, anemia is associated with a decrease in the supply of oxygen to damaged tissues, leading to poor wound healing[32]. Regarding the degree of glycemic control, the population was not similarly distributed in both groups, which does not allow an accurate analysis of the relationship of this variable with adverse outcomes. However, most patients with DFI had suboptimal glycemic control (HbA1c greater than 7% in 77.6% of patients with MDRB and 93.9% of non-MDRB patients), with an average HbA1c above 9%. In our study, there is also a high prevalence of anemia, which distorts the interpretation and influence of this variable.

### Importance in public health

Our study revealed a high rate of MDRB, much higher than most reports worldwide. This high rate could distort the impact on outcomes. Given this reality, it’s critical that all health facilities establish their bacteriological profile and susceptibility for DFI. This would facilitate the early and effective empirical use of antibiotics according to this profile, avoiding delays due to the use of limited utility antibiotics that could lead to infection progression. Many MDRB patients, due to intravenous treatments, require continued hospitalization, thereby increasing the intensive use of healthcare personnel. Extended stays heighten the risk of nosocomial infections, escalate direct costs, and limit the opportunity for other patients who require admission. The finding of an association with MDRB suggests an extreme need for careful biosecurity measures against cross-contamination and fomites from healthcare personnel and other patients.

### Limitations and Strengths

The high number of patients with MDRB could have limited their influence on outcomes in our cross-sectional analysis. We needed more non-MDRB patients to achieve a close-to-1 exposed/non-exposed ratio and improve estimation precision. Another limitation is the lack of cultures for anaerobic bacteria, which could have altered the distribution of germs in each group (MDRB and non-MDRB) when assessing the association. Additionally, our study’s findings cannot be generalized to other populations due to the presence of a Diabetic Foot program, which includes a multidisciplinary team that standardizes treatment, performing surgical debridement within the first 48 hours, contributing to the low rate of adverse DFI clinical outcomes observed.

Key strengths include our use of antibiotic therapy as per the IDSA guidelines and the procedures of the International Working Group of Diabetic Foot for managing peripheral arterial disease, offloading, glycemic management, and nutrition. Also noteworthy are our adherence to recommended standards for sample collection to avoid contamination and the use of a modern system for evaluating bacterial susceptibility.

## Conclusion

Our study did not find association between MDRB and death and/or major amputation, but it did with hospitalization exceeding 14 days. The high proportion of MDRB in the total sample and the cross-sectional design may have limited the discovery of differences in the main outcome. Further prospective studies, with larger sample sizes and equivalent studies, are needed to confirm its effect on these outcomes. The high prevalence of MDRB highlights the need for strategies to improve the timely identification of diabetic foot ulcers, as well as proper sample collection to identify pathogens and determine their susceptibility pattern to antibiotics before treatment initiation.

## Data Availability

All relevant data are within the manuscript and its Supporting Information files.

## Supporting information

**S1 Fig. Diagram of distribution of outcomes in the study’s patients**.

**S1 Table. Categories and agents used to define *Staphylococcus aureus* MDR, XDR, and PDR**.

**S2 Table. Categories and agents used to define *Enterococcus sp* MDR, XDR and PDR**.

**S3 Table. Categories and agents used to define *Enterobacteriaceae* MDR, XDR and PDR**.

**S4 Table. Categories and agents used to define *Pseudomonas aeuruginosa* MDR, XDR and PDR**.

**S5 Table. Categories and agents used to define *Acinetobacter spp* MDR, XDR and PDR**.

**S6 Table. Scales used in the study to assess the characteristics of the DFI. S7 Table. Data collection instrument**.

